# Lipid-lowering therapy for patients with arterial hypertension and concomitant chronic obstructive pulmonary disease in a south American cohort

**DOI:** 10.1101/2023.10.30.23297728

**Authors:** Diego Lobo, Santiago Bermudez, Luis Andrés Dulcey Sarmiento, Jaime Gomez, Carlos Hernandez, Juan Sebastián Theran Leon, Raimondo Caltagirone, Maria Camila Amaya, Maria Juliana Estévez Gómez, Diego Andrey Acevedo Peña, Silvia Fernanda Castillo Goyeneche, Anderson Felipe Arias, Edgar Blanco, Maria Ciliberti, Silvia Calderon, Emily Gutierrez, Angie Lizcano, Lizeth Bacca, Juan Camilo Martinez

## Abstract

**Introduction:** to assess the lipid-lowering effect, the effect on endothelial function, oxidative stress of Rosuvastatin at a dose of 40 mg in patients with dyslipidemia, arterial hypertension (AH) and chronic obstructive pulmonary disease (COPD) initially, over time after 4 weeks and 12 months of treatment.

**Material and methods:** The prospective study included 33 patients (mean age 60 [54;61] years) with hypertension, COPD and dyslipidemia. The laboratory examination consisted of determining the lipid spectrum and the level of lipid peroxidation products (LPO). To assess the tolerability of the prescribed therapy, creatinine, bilirubin, alanine aminotransferase (ALT), and aspartate aminotransferase (AST) were studied. To assess endothelial function, an endothelium-dependent vasodilation test (EDVD) was performed. Rosuvastatin at a dose of 40 mg was prescribed as lipid-lowering therapy. Initially, after 12 months, an ultrasound duplex scanning of the carotid arteries was performed to assess the presence of atherosclerotic plaques (AP) in the lumen of the vessel. The study was approved by the Local Ethics Committee of the Andes University in Merida, Venezuela (protocol No. 10 of December 25, 2017).

**Results:** after 4 weeks of treatment with Rosuvastatin at a dose of 40 mg, there was a significant decrease in total cholesterol levels by 26%, low-density lipoproteins (LDL) by 33%, triglycerides (TG) by 19%, while high-density lipoproteins (HDL) increased by 18%. Improvements in endothelial dysfunction (ED) and lipid peroxidation were observed during treatment with Rosuvastatin. Treatment with Rosuvastatin at a dose of 40 mg did not cause adverse reactions in patients.

**Conclusion:** correction of lipid metabolism disorders in patients with hypertension and COPD by prescribing Rosuvastatin at a dose of 40 mg can quickly reduce total cholesterol, LDL and TG, positively affecting endothelial function and lipid peroxidation processes. Therapy with Rosuvastatin at a starting dose of 40 mg in patients with dyslipidemia, hypertension and COPD is safe. After 12 months of regular use of Rosuvastatin at a dose of 40 mg, regression of AB was observed.

## INTRODUCTION

Among cardiovascular diseases, arterial hypertension (AH) is the most common, and some authors report that it coexists with chronic obstructive pulmonary disease (COPD) in 34% of patients [1–5]. Increased blood levels of proinflammatory cytokines (interleukin-8, interleukin-6, interleukin-1, and tumor necrosis factor α) are linked to extrapulmonary symptoms of COPD [1] [2]. Systemic inflammation, in turn, causes the arterial wall to become more stiff, especially in large-caliber arteries. This is a significant risk factor for cardiovascular problems [1] [2] [6]. Patients with COPD often have both localized bronchial inflammation and persistent systemic inflammation, which significantly contributes to the development of atherosclerosis in this patient population [1] [2]. As a result, individuals with COPD have a higher chance of experiencing an acute coronary event, particularly during exacerbations [1] [2] [7] [8]. This is because long-term systemic inflammation causes the coronary arteries to become more unstable due to the rapid development of atherosclerotic plaques, which also stiffen the arterial wall.

Endothelial dysfunction (ED) and oxidative stress are the foundations for the development and evolution of bronchopulmonary and cardiovascular diseases [1] [2], and these conditions start long before clinical ones manifest. Studying lipid peroxidation (LPO) and endothelial function is pertinent in this context since they are the first steps in a chain reaction of pathological responses that result in alterations to the vascular wall. It is indisputable and has been consistently demonstrated in numerous major trials [9–18] that hypertension patients with dyslipidemia should be prescribed lipid-lowering treatment, especially statins.

The usage of statins in COPD patients is an intriguing topic. A substantial body of research has been conducted demonstrating the potential benefits of this class of medications for both the progression of COPD and the prevention of cardiovascular illnesses [2]. By inhibiting inflammatory and remodeling effects, such as fibrosis processes, cytokine production, and neutrophil infiltration, statins act on the pathogenetic links in COPD [19–21]. As a result, a number of significant clinical effects are realized, such as a slowdown in the rate of decline in FEV1, a significant decrease in C reactive protein (CRP), a reduction in overall mortality, a reduction in COPD mortality, a reduction in the number of exacerbations, a decrease in hospitalizations, and increased exercise tolerance [19–21].

In patients with dyslipidemia, hypertension, and concomitant COPD, the study aimed to evaluate the hypolipidemic effect, the effect of 40 mg of rosuvastatin on endothelial function and oxidative stress, as well as the state of atherosclerotic plaques in the carotid arteries at the end of a 12-month treatment period. The Andes University’s Local Ethics Committee in Merida, Venezuela gave its approval to the study (protocol No. 10 of December 25, 2017).

## MATERIAL AND METHODS

The prospective study included 33 patients with hypertension, COPD and dyslipidemia (Table 1). The average age of the patients was 60 [54;61] years.

**Table 1.** General characteristics of patients participating in the study.

| Index | n =33 |
| --- | --- |
| Men, abs. (% in Group) | 15 (45.45%) |
| Women, abs. (% in Group) | 18 (54.55%) |
| Average age, years | 60 [54;61] |
| Duration of hypertension, years | 6 [4;7] |
| Duration of COPD, years | 14 [11;15] |
| AG 1st century, abs. (% in Group) | 12 (36.38%) |
| AG 2 tbsp., abs. (% in Group) | 19 (57.56%) |
| AG 3 st., abs. (% in Group) | 2 (6.06%) |
| IR, packs/years | 20 [14;24] |
| COPD according to GOLD 1st degree, abs.<br>(% in Group) | 15 (45.44%) |
| COPD according to GOLD, grade 2, abs.<br>(% in Group) | 8 (24.26%) |
| COPD according to GOLD, grade 3, abs.<br>(% in Group) | 10 (30.3%) |
| CAT < 10, abs. (% in Group) | 21 (63.66%) |
| CAT >10, abs. (% in Group) | 12 (36.34%) |
| mMRc < 2, abs. (% in Group) | 24 (72.77%) |
| mMRc > 2, abs. (% in Group) | 9 (27.23%) |
| Moderate SVR (SCORE scale) | 4 (12.12%) |
| High SSR (SCORE scale) | 22 (66.67%) |
| Very high CCP (SCORE scale) | 7 (21.21%) |
| Total cholesterol, mmol/l | 6.11 [5.9;6.5] |
| LDL, mmol/l | 4.12 [3.9;4.6] |
| HDL, mmol/l | 0.98 [0.9;1.1] |
| TG, mmol/l | 3.1 [2.8;3.6] |
| Bilirubin, μmol/l | 9.8 [6.9;12.2] |
| AST, U/l | 21.5 [19.6;28.0] |
| ALT, U/l | 24.0 [19.0;27.4] |
| EDVD, % | 4.9 [4.5;5.5] |
| Imax/S | 0.22 [0.19;0.26] |
| OSH/(DK+TK) | 78.2 [71.27;84.49] |

Note: data are presented as median and interquartile range (Me [25r; 75r] ) and the absolute number of patients (% of the total). Analysis of the type of distribution was assessed by the Kolmogorov-Smirnov test.

Ages over eighteen, essential arterial hypertension, GOLD-rated COPD grades 1-3, without exacerbation, laboratory-confirmed dyslipidemia, and voluntary informed consent to participate in the study were the inclusion criteria for the research. Secondary forms of arterial hypertension, CHF III–IV FC, chronic heart failure (CHF) with reduced ejection fraction, valvular heart disease with hemodynamically significant disorders, COPD exacerbation, oncological diseases, pregnancy, lactation, bronchial asthma, and patients who experienced acute inflammatory diseases within a month prior to the study’s commencement were among the exclusion criteria.

According to the 2018 European guidelines and the 2020 International guidelines for hypertension, patients were first treated with antihypertensive therapy using a variety of classes of antihypertensive medications, such as beta-blockers, ACE inhibitors, diuretics, and angiotensin II receptor blockers. After reaching target blood pressure levels of less than 130/80 mmHg, patients were added to the study. [4] [5] Art. In accordance with the 2019 GOLD clinical guidelines, beta2-agonists, M-anticholinergics, inhaled glucocorticosteroids, or a mix of these bronchodilator classes were used to treat COPD [7]. The bronchodilator doses were not altered during the study because the patients were not in the acute stage.

The criteria outlined in the clinical guidelines for hypertension (ESC/ECH 2018, RSC 2020) [4] [5] were used to make the diagnosis of hypertension. The patient’s blood pressure reading prior to antihypertensive medication prescription was used to assess the patient’s degree of hypertension. The SCORE scale was used to calculate the risk of cardiovascular complications. The Global Initiative for COPD (GOLD, 2017, 2020) criteria were used to establish the diagnosis of COPD. The most recent European clinical guidelines on dyslipidemia from 2019 were used to define the criteria for dyslipidemia [22].

A general examination of the organs and systems, anamnestic data collection, supplementary laboratory and instrumental studies, and a patient’s complaints were all part of the examination process.

The laboratory examination comprised measuring the levels of primary (diene conjugate (DC), secondary (triene conjugate (TK)), and final products of LPO (Schiff bases (OS)) using the method of I.A. Volchegorsky (1989), as well as the lipid spectrum (total cholesterol (TC), low-density lipoproteins (LDL), high-density lipoproteins (HDL), triglycerides (TG), and others. Additionally, the intensity of free radical oxidation was measured using the method of E.I. Kuzmina, A.S. Nelyubina, and M.K. Shchennikova, 1983 (S, Imax). An analysis was conducted on creatinine, bilirubin, alanine aminotransferase (ALT), and aspartate aminotransferase (AST) to evaluate the long-term tolerability of the recommended therapy.

A test using endothelium-dependent vasodilation (EDVD) and ultrasound duplex scanning of the carotid arteries (USDS) were two examples of instrumental research methods. Celermajer D. (1992) states that a high-resolution ultrasound sensor (HRUS) was used in a non-invasive ultrasound method to assess endothelial dysfunction. To find the presence of atherosclerotic plaque (AB), ultrasound duplex scanning of the carotid arteries (CA) was carried out using a Vivid-7 device with a linear sensor 9–11 MHz. Local thickening of the SA area by more than 0.5 mm or 50% in comparison with surrounding areas, or thickening of the SA area by more than 1.3 mm with its protrusion towards the vessel lumen, were the criteria for the presence of AB in the SA [4] [5]. At the location of the AB localization, which matched the ECST method (The European Carotid Surgery Trial), the percentage of stenosis was directly measured. The maximum percentage of stenosis in a specific patient (MaxStSA) was also calculated, as well as the total value of carotid artery stenosis (SumStSA), which is the sum of the percentages of all carotid artery stenoses on both sides.

As a lipid-lowering medication, 40 mg of rosuvastatin (Livazo, Recordati, Ireland) was prescribed to each patient. Initial therapy with a dose of 40 mg is safe and highly effective, according to the results of the international observational program LEADER, which compared different treatment regimens with Rosuvastatin, namely 1 mg, 2 mg, and 40 mg in patients with high and very high cardiovascular risk [23]. Clinical guidelines [22] state that the objective of lipid-modifying therapy should be to achieve target values, which include lowering LDL cholesterol levels consistently to levels seen in the most recent large-scale PCSK9 inhibitor studies in order to reduce the risk of atherosclerosis. Accordingly, it is advised that patients who are considered to be at very high risk have their LDL cholesterol levels reduced by at least 50% from baseline for both primary and secondary prevention, with the goal of achieving a level of less than 1.4 mmol/L (less than 55 mg/dL). It is advised that LDL cholesterol levels in patients at high cardiovascular risk be lowered by at least 50% from baseline in order to reach the desired level of less than 1.8 mmol/L (less than 70 mg/dL). LDL-C targets of <2.6 mmol/L (<100 mg/dL) are advised for patients at moderate risk, and <3.0 mmol/L (<116 mg/dL) for patients at low risk [22].

STATISTICA 10.0, a licensed program, was used to process statistical data. Kolmogorov-Smirnov test was used to analyze the distribution type. The Wilcoxon test was used to compare two dependent groups, specifically to ascertain the safety and efficacy of treatment between visits within the group. The Kruskal-Wallis test, a nonparametric technique, was used to determine the statistical significance of the differences between the three groups, and pairwise comparison was then performed. When comparing three groups pairwise, differences were deemed statistically significant at p<0.05 and p<0.017 (using the Bonferroni correction).

## RESULTS

After 4 weeks of treatment with Rosuvastatin at a starting dose of 40 mg, there was a significant decrease in all indicators characterizing the lipid spectrum, namely total cholesterol decreased by 26% from the original, LDL - by 33%, TG - by 19%, while HDL increased by 18% from the original (Table 2).

**Table 2.** Dynamics of lipid spectrum indicators, as well as bilirubin, AST and ALT during treatment with Rosuvastatin 40 mg.

| Index | Originally | In 4 weeks | p value |
| --- | --- | --- | --- |
| Total cholesterol, mmol/l | 6.2 [5.9;6.5] | 4.6 [4.3;4.9] | $p < 0.001$ |
| LDL, mmol/l | 4.2 [3.9;4.2] | 2.8 [2.8;3.1] | $p < 0.001$ |
| HDL, mmol/l | 0.9 [0.9;1.4] | 1.1 [0.9;1.1] | $p < 0.001$ |
| TG, mmol/l | 3.1 [2.9;3.6] | 2.5 [2.3;2.8] | $p < 0.001$ |
| Creatinine, $\mu\text{mol/l}$ | 124.3 [96.8;129.2] | 129.3 [127.3;140.4] | $p = 0.09$ |
| Bilirubin, $\mu\text{mol/l}$ | 9.8 [6.9;12.2] | 9.3 [7.1;10.4] | $p = 0.15$ |
| AST, U/l | 21.5 [19.6;28.0] | 21.0 [18.6;24.2] | $p = 0.35$ |
| ALT, U/l | 24.0 [19.0;27.4] | 21.2 [18.3;24.0] | p=0.14 |

**Note:** data are presented as median and interquartile range (Me [25p; 75p] ), Wilcoxon test. Differences were considered statistically significant at $p < 0.05$ .

Initially, the primary products of LPO, namely DC and TC, as well as the final products of LPO in the form of OR, exceeded the norm. The analysis carried out using the nonparametric Kruskal-Wallis method showed that the accumulation of lipid peroxidation products and the level of ET-1 increases both with the increase in the severity of obstructive disorders of COPD and cardiovascular risk (Table 3). It should be noted that among the studied indicators of ED, only for the ET-1 level, a dependence was revealed on the degree of bronchial obstruction (Kruskal-Wallis test = 14.7; p < 0.001) and cardiovascular risk (Kruskal-Wallis test = 34.76; p < 0.001).

**Table 3.**
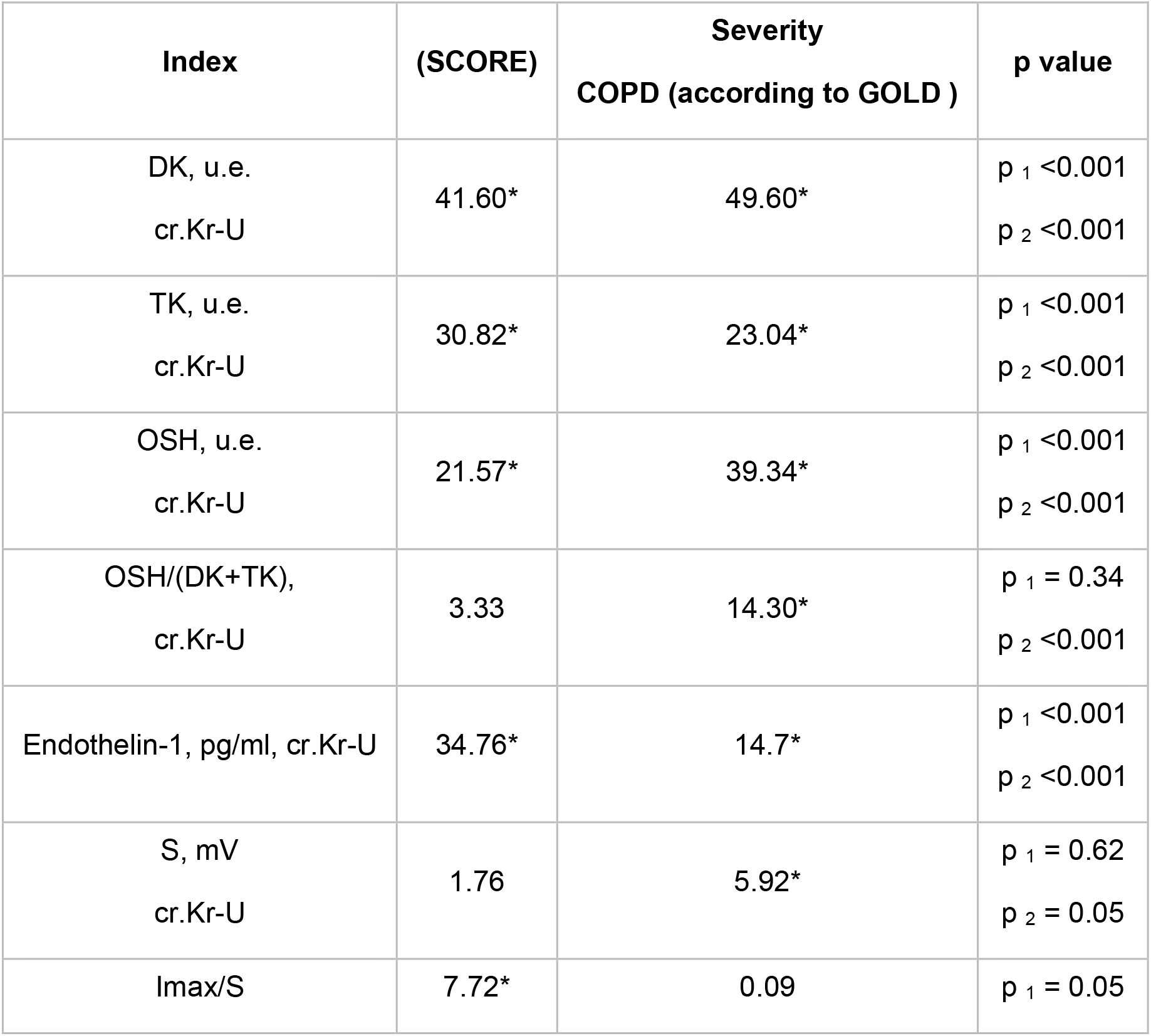
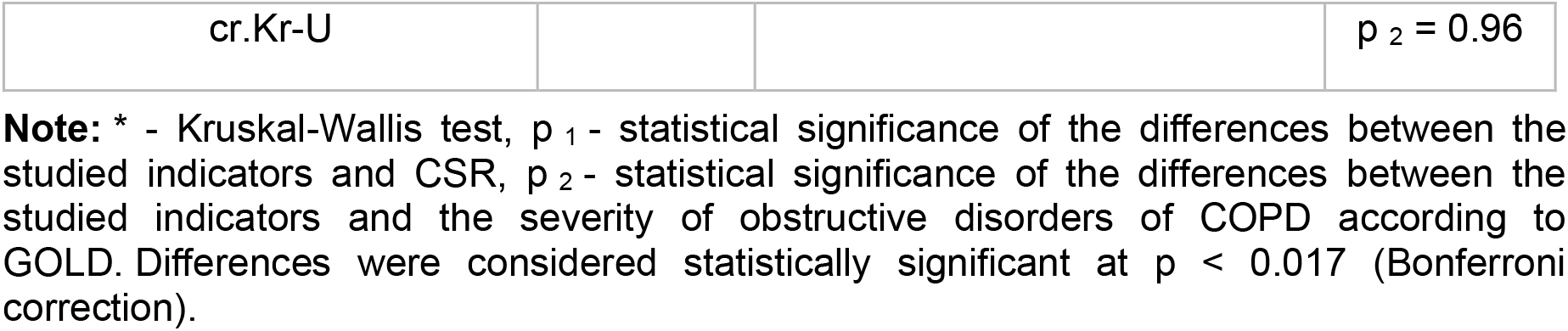
Dependence of indicators of oxidative stress and endothelial dysfunction on the severity of COPD and cardiovascular risk.

During treatment with Rosuvastatin, after 4 weeks of treatment, an improvement in ED indicators and lipid peroxidation processes was observed (Table 4). Statistically significant results were obtained for DC, TC, OR, S, Imax. A significant increase in the ratio OR/(DC+TC) after 4 weeks of treatment is associated with a decrease in primary products (PC) and accumulation of end products of lipid peroxidation (OR). When assessing the indicators of the antioxidant defense system (Imax), their normalization was noted.

**Table 4.**
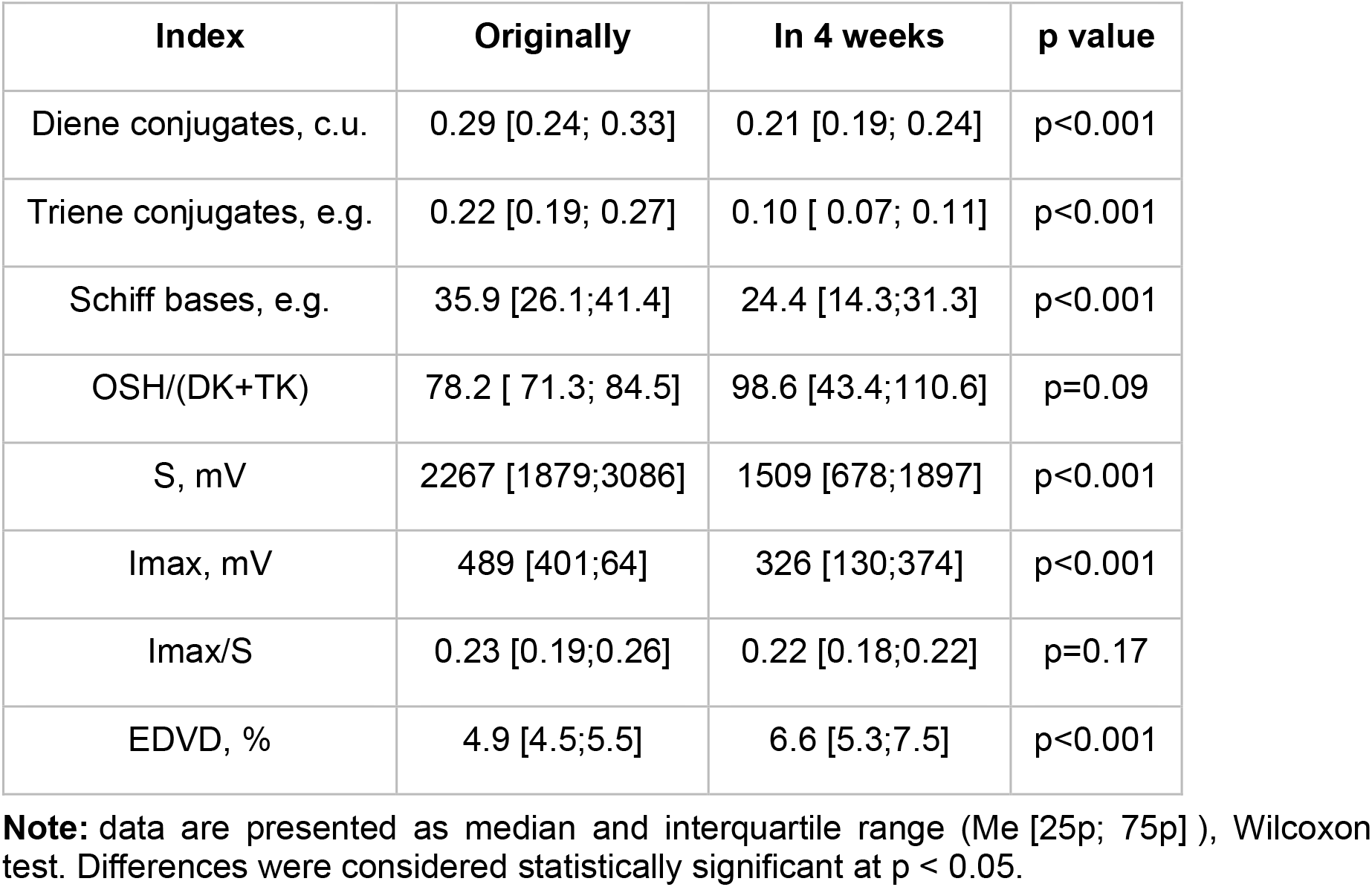
Dynamics of indicators of oxidative stress and endothelial function during treatment with Rosuvastatin 40 mg.

| Index | Originally | In 4 weeks | p value |
| --- | --- | --- | --- |
| Diene conjugates, c.u. | 0.29 [0.24; 0.33] | 0.21 [0.19; 0.24] | $p < 0.001$ |
| Triene conjugates, e.g. | 0.22 [0.19; 0.27] | 0.10 [0.07; 0.11] | $p < 0.001$ |
| Schiff bases, e.g. | 35.9 [26.1; 41.4] | 24.4 [14.3; 31.3] | $p < 0.001$ |
| OSH/(DK+TK) | 78.2 [71.3; 84.5] | 98.6 [43.4; 110.6] | $p = 0.09$ |
| S, mV | 2267 [1879; 3086] | 1509 [678; 1897] | $p < 0.001$ |
| I<sub>max</sub>, mV | 489 [401; 64] | 326 [130; 374] | $p < 0.001$ |
| I<sub>max</sub>/S | 0.23 [0.19; 0.26] | 0.22 [0.18; 0.22] | $p = 0.17$ |
| EDVD, % | 4.9 [4.5; 5.5] | 6.6 [5.3; 7.5] | $p < 0.001$ |

**Note:** data are presented as median and interquartile range (Me [25p; 75p]), Wilcoxon test. Differences were considered statistically significant at $p < 0.05$ .

Initially, when performing a test with EDVD, patients observed a decrease in the diameter and speed of blood flow in the brachial artery, while EDVD was 4.9 [4.5;5.5] %. After 4 weeks of treatment, there was a statistically significant increase in EDVD to 6.6 [5.3;7.5] %. It should also be noted that treatment with Rosuvastatin at a dose of 40 mg did not cause any adverse reactions in patients, and there was no increase in creatinine, bilirubin, AST and ALT, which indicates the safety of using this drug, including at a high dose of 40 mg (Table 2).

Ultrasound duplex scanning of the carotid arteries, as one of the basins of subclinical course of atherosclerosis, most accessible for visualization using ultrasound, was carried out after 12 months of treatment. A limitation of this stage of this study was the lack of data on the state of the lipid spectrum and other indicators performed over time after 4 weeks of observation in most patients, which did not allow them to be analyzed after 12 months of treatment. However, the value of information that allows us to assess the status of identified atherosclerotic plaques in the carotid arteries, which would be illogical to do after 4 weeks or another shorter period of time due to the lack of rapid dynamics of these data, prompted us to analyze the ultrasound data of the carotid arteries at baseline and after 12 months treatment with Rosuvastatin at a dose of 40 mg. Available and at the same time generally accepted indicators for assessing atherosclerotic plaques, such as SumStCA and MaxStCA, were analyzed. Ultrasound scanning of the carotid arteries revealed AB in 29 (87%) patients included in the study. After 12 months of treatment with Rosuvastatin 40 mg, SumStSA decreased by 7.5% (from 40.2% to 37.2%), and MaxStSA decreased by 4.6% (from 49.5% to 47.2%).

## DISCUSSION

Regardless of smoking status, concurrent COPD in patients with cardiovascular pathology can appropriately be regarded as an extra cardiovascular risk factor. Proinflammatory cytokine activation in COPD sets off a series of pathological events that result in endothelium dysfunction, an imbalance in the oxidant-antioxidant system, and an early, aggressive course of atherosclerosis.

Significant abnormalities in lipid peroxidation, eDVD, and lipid spectrum were found in our study’s patients with COPD and hypertension, necessitating extremely intensive lipid-lowering therapy. 40 mg of rosuvastatin was the starting dosage that was prescribed.

Our research demonstrated that, after just 4 weeks of treatment, rosuvastatin given at a high starting dose of 40 mg can reduce lipid peroxidation, activate the antioxidant defense system, and improve endothelial function—all of which will undoubtedly stop atherosclerosis from progressing. Lipid profiles improved significantly after 4 weeks of treatment, but LDL target values were not reached, which is not uncommon in clinical practice, particularly in patients with high and very high cardiovascular risk as determined by SCORE. This is also in line with the most recent recommendations regarding dyslipidemias [22], necessitates taking a second medication to lower cholesterol. Notably, despite the high dosage of 40 mg of rosuvastatin, no adverse effects were noted, and there was no decline in the biochemical parameters of the liver and kidneys.

Thus, we can conclude that initial therapy with 40 mg of rosuvastatin is safe and highly effective based on our results as well as the results of the international observational program LEADER [23].

Numerous studies (the Japan Assessment of Rosuvastatin and Atorvastatin in Acute Coronary Syndrome study, TOGETHAR trial) that used intravascular ultrasound for analysis and showed the effect of rosuvastatin on atherosclerosis through stabilization and regression of AB suggest a high accuracy of the results obtained. Our study was limited by the use of carotid artery ultrasonography, which is less accurate but is frequently utilized in clinical settings and might therefore be of use to practitioners. The production of apoA1, the expression of scavenger receptor B1 in macrophages, and the stimulation of cholesterol exchange between cells and HDL are some of the mechanisms of AB regression associated with rosuvastatin use [17]. Following a year of 40 mg of rosuvastatin treatment, we saw a regression of arterial hypertension in the carotid arteries. This suggests that rosuvastatin can slow down the rate at which atherosclerosis progresses, which in turn causes the atherosclerotic process to regress, thereby lowering the risk of cardiovascular complications indirectly.

## CONCLUSION

When patients with concomitant COPD and hypertension have their lipid metabolism disorders corrected, rosuvastatin at a starting dose of 40 mg can quickly lower total cholesterol, LDL, and TG while improving endothelial function and oxidative stress. It is safe to start rosuvastatin therapy at 40 mg for patients with COPD, hypertension, and dyslipidemia. Regression of AB volume was seen after a year of consistent 40 mg doses of rosuvastatin use.

## Data Availability

All data produced in the present work are contained in the manuscript

## Financing

The study had no sponsorship.

